# Prophylactic Magnesium Supplementation and New-Onset Atrial Fibrillation in a General Critical Care Population: A Prescribing Preference Instrumental Variable Analysis

**DOI:** 10.1101/2021.11.25.21266861

**Authors:** Matthew G. Wilson, Aasiyah Rashan, Roman Klapaukh, Folkert W. Asselbergs, Stephen K. Harris

**Author notes:** Corresponding author; @MWilson1987. These authors contributed equally to this work (Matthew G. Wilson, Aasiyah Rashan).

## Abstract

**Aims:** Atrial fibrillation is frequently encountered in critical illness and causes adverse effects including haemodynamic decompensation, stroke and longer hospital stay. It is common to supplement serum magnesium for the purpose of preventing new-onset atrial fibrillation. However, no randomised studies support this practice in the non-cardiac surgery critical care population, and its effectiveness is unclear. We sought to investigate the effectiveness of magnesium supplementation in preventing new-onset atrial fibrillation in a mixed critical care population.

**Methods:** We conducted a single centre retrospective observational study of adult critical care patients. We employed a natural experiment design, using the supplementation preference of the bedside critical care nurse as an instrumental variable.

Using the electronic patient record, magnesium supplementation opportunities were defined and linked to the bedside nurse. Nurse preference for administering magnesium was obtained using multilevel modelling. The results were used to define ‘pro’ and ‘anti’ supplementation groups, which were inputted into an instrumental variable regression to obtain an estimate of the effect of magnesium supplementation.

**Results:** 9,114 magnesium supplementation opportunities were analysed, representing 2,137 critical care admissions for 1,914 patients. There was significant variation in magnesium supplementation practices attributable to the individual nurse, after accounting for covariates. The instrumental variable analysis showed magnesium supplementation was associated with a 3% decreased chance of experiencing new-onset atrial fibrillation (95% CI −0.06 to −0.04, p = 0.03).

**Conclusions:** This study supports the strategy of routine magnesium supplementation, but further work is required to identify optimal serum magnesium targets for prophylaxis of atrial fibrillation.

**What’s New?:**

- Routine administration of supplemental magnesium sulphate is associated with a reduced chance of developing new-onset Atrial Fibrillation, in a general critical care cohort.
- This finding agrees with previous Randomised Controlled Trial results which are limited to the cardiac critical care population.
- This finding disagrees with previously published observational studies, which is likely due to better control of unmeasured confounding.
- There is significant variation in serum magnesium supplementation attributable to the individual bedside critical care nurse.
- Electronic health records offer the ability to evaluate the effectiveness of routinely administered treatments which lack evidence in an affordable way.
- Natural experiments and instrumental variable analysis offer the opportunity to derive causally robust estimations of treatment effectiveness by better accounting for unobserved confounding.

## Introduction

New-Onset Atrial Fibrillation (NOAF) is a common accompaniment to critical illness. Together with pre-existing atrial fibrillation, it is observed in nearly one-third of patients passing through critical care^1^. The incidence of NOAF is highest following cardiac surgery (25-50%), major thoracic surgery and oesophagectomy (10-30%), and is less common following extra-thoracic surgery (11%) ^2–5^. In contrast, medical CCU patients have a general incidence of 11%, with high-risk groups (e.g., septic shock) exhibiting a prevalence of 42% in one study ^6^.

NOAF arises from disturbance to the balance of pro-arrhythmogenic factors and opposing compensatory mechanisms. Bosch et al have described in detail the physiological processes underpinning the development of “arrhythmogenic atria” in the context of critical illness. Changes to electrolyte concentration can often provide the trigger to atria which have been “primed” to become arrhythmic ^1^.

Magnesium (Mg) plays a key role in regulating the cardiac action potential, through its action as a co-factor at the Sodium/Potassium ATPase pump, and at specific ion channels e.g., L-type Calcium channels. Mg offers cell membrane stabilising properties which may help maintain sinus rhythm in “at risk” atria ^7^.

Although hypomagnesaemia (serum Mg < 0.6 mmol/L) is common in critical care, the evidence supporting supplementation is mixed. The bulk of evidence comes from cardiac surgery, with the two most recent meta-analyses of existing Randomised Controlled Trials (RCTs) and a Cochrane Review finding supplemental Mg (or a higher serum Mg level) conveys a protective effect against developing NOAF ^8–10^. This association has not been universally replicated in observational work, with several studies reporting the opposite relationship ^11–13^. It is possible, that any inverse relationship seen in observational studies is a consequence of confounding by indication: that patients at greater risk of NOAF are more likely to receive supplementation.

Overall, outside the cardiac surgical population, there is little evidence supporting the routine supplementation of Mg for the purpose of preventing NOAF. Nevertheless, Mg administration, even at normal serum Mg levels, continues to form a routine part of critical care.

There are currently seven clinical trials of Mg for atrial fibrillation registered on clinicaltrials.gov (*June, 2021*). Of these, two are recruiting medical patients, the remainder focus on cardiac surgery. No studies address the question of the efficacy of Mg supplementation for NOAF *prophylaxis*, instead focusing on its use as a treatment strategy.

Given the difficulties inherent to using observational methods to derive estimates of treatment effects, quasi-experimental techniques may offer potential solutions. Finding opportunities to conduct natural experiments can help limit the confounding effects of selection bias, and confounding by indication. In particular, natural experiments based on clinician prescribing preferences have been used to successfully investigate the efficacy of treatments such as non-steroidal anti-inflammatories and emergency general surgery ^14, 15^.

In this study, we identified a natural experiment to evaluate the effect of Mg supplementation on developing NOAF. Specifically, that the bedside critical care nurse’s preference for Mg administration under varying conditions, may act as an instrumental variable to derive a more causally robust treatment effect estimate. We categorised individual nurses into ‘liberal’ or ‘restrictive’ Mg supplementation groups, based on their observed behaviour. That is, given the same measured serum Mg, are they more or less likely to supplement, relative to their colleagues in similar situations. Having dichotomised the nurse population according to prescribing preference, we then evaluated the relationship between patient exposure to liberal or restrictive nurse (the instrument) and the subsequent incidence of NOAF. This permitted us to ascertain a more robust estimate of the association, by accounting for unmeasured confounding.

## Methods

### Ethics and Data Governance

This study used routinely collected Electronic Health Record (EHR) data from critical care admissions to a tertiary referral centre in the United Kingdom. Patient data was collected under the auspices of the Critical Care Health Informatics Collaborative (CCHIC). CCHIC has an exemption to collect identifiable clinical data under an opt-out consent framework, for the purposes of conducting secondary research. This is facilitated through an exemption to standard confidentiality law, detailed under Section 251 of the National Health Service (NHS) Act, 2006 (14/CAG/1001) and approved by the National Research Ethics Service (14/LO/103) ^16^. An application for data access pertinent to the study question was approved by the CCHIC Scientific Advisory Group. Data was stored and analysed in the University College London Data Safe Haven, a secure data environment conforming to NHS Digital’s Information Governance Toolkit ^16^.

### Study Population

All adult admissions to University College London Hospital (UCLH) 35-bed general adult critical care unit between January 2016 and December 2017 were examined. Critical care patients have a daily serum Mg measurement taken, together with other routine blood tests. An “as required” prescription for supplemental Mg is available for the bedside nurse to access and administer as they see fit.

For the purposes of the study, each patient’s admission was divided into ‘Observation Windows’, roughly equivalent to the nursing day shift. These windows consisted of a serum Mg measurement available at 8am, a subsequent opportunity for supplementation associated with the bedside ICU nurse, and a period of observation for NOAF prior to the next serum Mg measurement. Each patient therefore contributed an observation for as many days as they had serum Mg measured.

### Data Preparation

In detail, each observation window starts with the measured serum Mg date-time stamp. Observation windows were censored by the earliest of the next serum Mg measurement, the end of the patient episode (discharge from ICU), or 24 hours following initial measurement.

To reliably pair an individual CCU nurse with the observed serum Mg measurement, only serum Mg measurements recorded between midnight and 8am were included. Thus, in the final data table, each row represents a serum Mg measurement which may be observed by the bedside nurse during the day shift. Similarly, Mg administrations occurring after 8pm were removed as these would be associated with a different nurse.

All the recorded values for heart rhythm were joined to the relevant observation windows using date-time stamps. Heart rhythm data is recorded as 31 different categories within CCHIC. These were condensed to a binary indication of sinus rhythm or AF. The AF category comprised atrial flutter, atrial tachycardia, supraventricular tachycardia, and supraventricular tachycardia with aberrant conduction.

Using the same process, covariates deemed to be clinically relevant, and which are routinely collected by CCHIC, were extracted and assigned to the relevant observation window. Included covariates and transformations are listed in **Supplementary Table S1**, with an example of the observation windows in **Supplementary Table S2**.

NOAF was defined as a change in documented heart rhythm from the sinus rhythm category to the AF category, occurring within the observation window. As part of the patient’s routine observations on the CCU, the bedside nurse inputs the current heart rhythm at a minimum hourly cadence from direct observation of the continuous cardiac monitor.

### Exclusion Criteria

Observation windows were excluded from the analysis if the patient was already in atrial fibrillation (not new-onset), or where the Mg was administered after a NOAF event (indicating administration for treatment, rather than prophylaxis). As complete cases are required for an instrumental variable analysis, observation windows missing covariate data were excluded. Overall, this represented 3% (512) of the total number of observation windows examined. A breakdown of missing data is illustrated in **Figure 1**. Infusions such as noradrenaline were assumed to be switched off rather than missing if infusion rate was not reported. This matches clinical reporting practice on the CCU.

**Figure 1.**
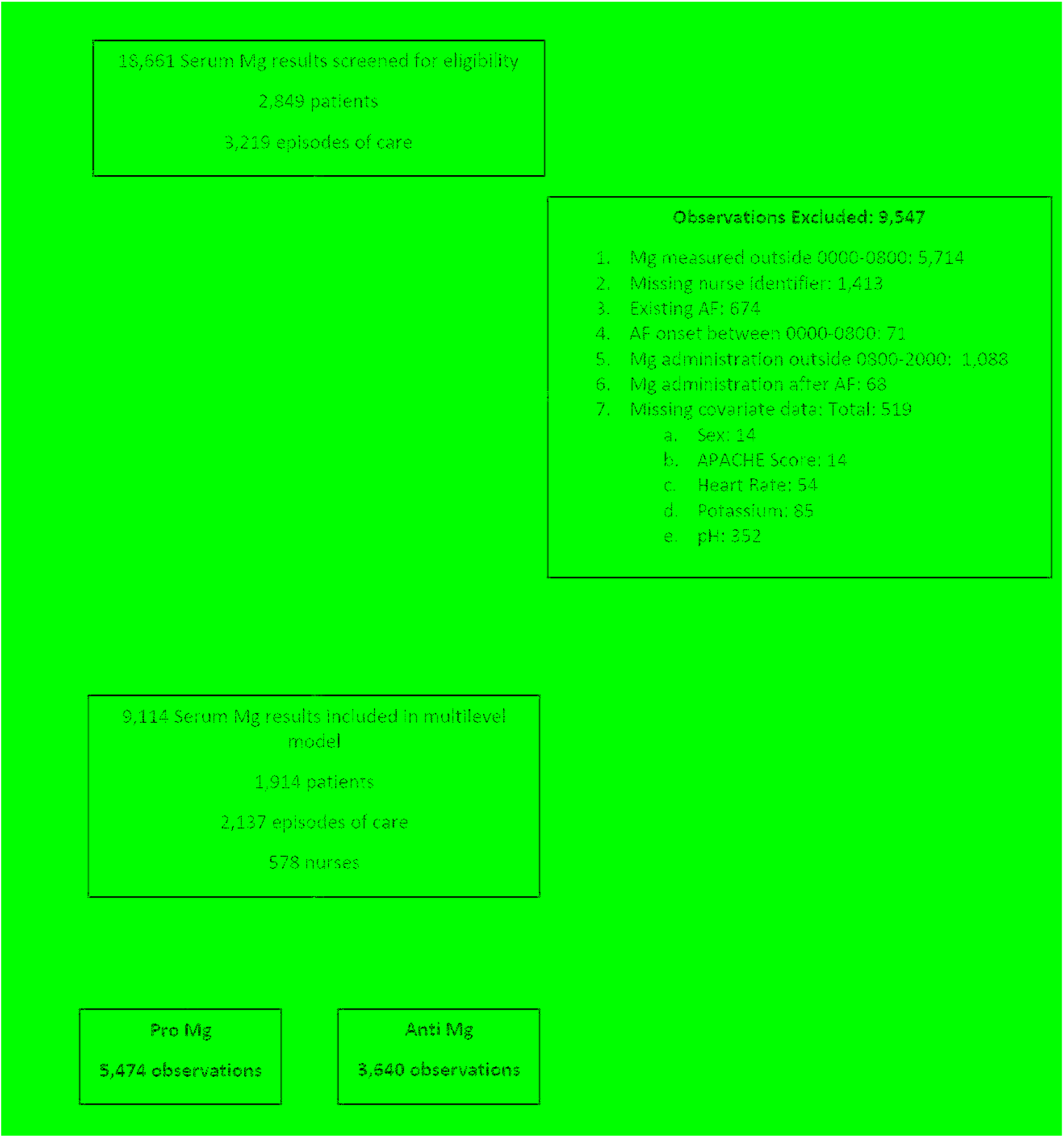
Study CONSORT Diagram.

### Statistical Analysis

To identify the extent of variation in Mg supplementation practices and thus establish individual nurse preferences, we constructed a multilevel model to predict the likelihood of supplementation for each observation window. Windows were nested by nurse identity, which acted as a random effect in the model. Additional covariates which were deemed to be clinically relevant to the supplementation decision were added to the model as fixed effects. These included measured serum Mg, previous atrial fibrillation within the same critical care admission, noradrenaline dose, and illness severity (Acute Physiology and Chronic Health Evaluation II score). Individual covariates were added in a step-wise fashion to optimise inclusion of clinically important variables, minimise the model Akaike Information Criteria value, and maintain model convergence.

The predicted probabilities of Mg supplementation for each observation window, nested in individual critical care nurse identities were then used to conduct a prescribing preference instrumental variable analysis. In keeping with this design, it was assumed that the allocation of nurse to a particular patient on a particular day was both random in nature, and could not affect that patient’s chance of developing NOAF, other than through the nurse’s preference for supplementing Mg. A detailed discussion of the assumptions required by the instrumental variable analysis is available in the supplementary materials. Keele et al present a detailed explanation of this method in their work on estimating the effect of emergency general surgery ^15^.

Nurse identities were divided into ‘Liberal’ and ‘Restrictive’ Mg supplementation groups by splitting the random intercepts assigned to each nurse by the median intercept value obtained from the multilevel model. Following this, a two-stage least squares (2SLS) instrumental variable regression model was constructed to estimate the effect of Mg supplementation on NOAF occurrence within each observation window, using the nurse’s group assignment as the instrument. Clinically relevant covariates were then added. The instrumental variable model was tested using the weak instrument F test on the nurse group ^17^, and the Wu-Hausman test for endogeneity ^18^. To address potential violation of the 2SLS linearity assumption, a further instrumental variable model was constructed, using a Probit link to account for the binary outcome variable. We performed data cleaning, multilevel modelling and 2SLS regression in R ^19^. We performed the instrumental variable Probit analysis in Stata ^20^.

## Results

9,114 observation windows were included in the analysis, representing 2,137 critical care admissions for 1,914 patients. **Figure 1** illustrates participant flow through the study, with data loss at each stage. There were 578 nurses associated with the observation windows. **Table 1** summarises the patient characteristics for the sample population. The mean serum Mg on admission to critical care was 0.94 mmol/L (SD 0.24 mmol/L). 55% (1,057) of patients received at least one Mg supplementation during their admission. 5.38% (103) of patients had at least one documented episode of NOAF.

**Table 1.** Summary of Study Population

| Variable | Summary |
| --- | --- |
| Age (Years)* | 62 (24) |
| Sex n (%) | 979 (51.2) Female |
| Length of Critical Care Stay (Days)* | 3.58 (5.08) |
| APACHE II Score* | 17 (9) |
| Serum potassium on admission (mmol/L)** | 4.51 (0.56) |
| Serum magnesium on admission (mmol/L)** | 0.97 (0.24) |
| Serum pH on admission** | 7.39 (0.06) |
| At least one magnesium supplementation n (%) | 1,057 (55.22) |
| At least one atrial fibrillation event n(%) | 103 (5.38) |

\*Non-normally distributed variables described using median (interquartile range);
\*\*Normally distributed variables described using mean (standard deviation)

The first stage analysis constructed a multilevel model to estimate the probability of Mg supplementation in each observation window, nested by individual critical care nurse. The results from this model are included in **Supplementary Table S3**. After adjusting for measured serum Mg, previous atrial fibrillation within the same critical care admission, noradrenaline dose and illness severity, approximately 32% of variation in Mg supplementation observed in the model was attributable to the individual nurse.

Using the multilevel model, predicted probabilities for Mg supplementation within each observation window were obtained. These were used to divide nurses into ‘Liberal’ and ‘Restrictive’ supplementation groups for the instrumental variable analysis. 289 nurses were assigned to each group, corresponding to 5,474 observation windows in the ‘Liberal’ group and 3,640 in the ‘Restrictive’ group. Nurses in the ‘Liberal’ group observed a median of seven observation windows (IQR 32), and nurses in the ‘Restrictive’ group observed a median of three (IQR 17) observation windows.

**Tables 2** and **3** illustrate the characteristics of individual patients, and observation windows (the unit of analysis), in both supplementation groups. Overall, both groups were similar for all the included covariates, but differed significantly in Mg supplementation (2,640 administrations in the ‘Liberal’ group, versus 1,027 administrations in the ‘Restrictive’ group). The incidence of NOAF was higher (78 episodes, 2.03%) in the ‘Restrictive’ supplementation group compared to the ‘Liberal’ group (74 episodes, 1.74%).

**Table 2.** Patient Level Characteristics of Liberal and Restrictive Magnesium Supplementation Groups

| Variable | Liberal | Restrictive |
| --- | --- | --- |
| Age (Years), Median (IQR) | 62 (23) | 61 (24) |
| Sex (%) | Female: 50 | Female: 51 |
| Length of Stay (Days), Median (IQR) | 4.4 (5.9) | 4.7 (6.7) |
| APACHE II Score, Median (IQR) | 17 (9) | 17 (9) |
| Unit Mortality (%) | Dead: 10.9 | Dead: 11.8 |

**Table 3.** Observation Window Characteristics of Liberal and Restrictive Magnesium Supplementation Groups

| Variable | Liberal (n = 5,474) | Restrictive (n = 3,640) |
| --- | --- | --- |
| Mean (SD) Serum potassium (mmol/L) | 4.58 (0.50) | 4.58 (0.51) |
| Mean (SD) Serum magnesium (mmol/L) | 0.94 (0.21) | 0.95 (0.21) |
| Mean (SD) Serum pH | 7.40 (0.06) | 7.40 (0.06) |
| Mean (SD) Noradrenaline use (prev. 24 hours) (mcg.kg.min<sup>-1</sup>) | 0.02 (0.07) | 0.02 (0.07) |
| Number of magnesium administrations (%) | 2,640 (48.2) | 1,027 (28.2) |
| Number of atrial fibrillation events (%) | 78 (1.42) | 74 (2.03) |

The results of the instrumental variable regression estimating the association between Mg supplementation and NOAF are summarised in **Table 4**. After accounting for age, sex, illness severity (APACHE II Score), previous atrial fibrillation during the same CCU admission, baseline serum Mg, serum potassium, pH, heart rate, and mean noradrenaline consumption, Mg supplementation was associated with a 3% decrease in chance of experiencing NOAF (95% CI −0.06 to −0.004, p = 0.03).

**Table 4.**
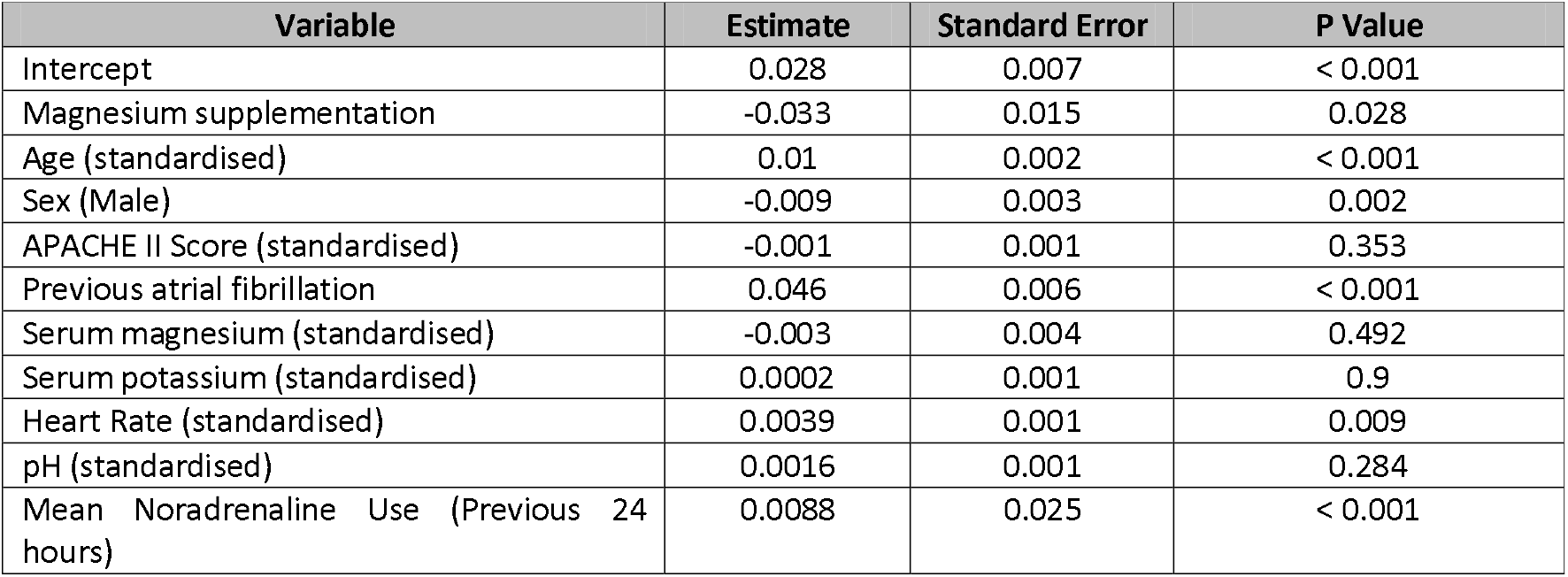
Effect of Magnesium Supplementation on Chance of New-Onset Atrial Fibrillation

The weak instrument test indicated that nurse group was predictive of receiving Mg supplementation (p < 0.01), and was therefore not a weak instrument. The Wu-Hausman test suggested the presence of endogeneity in the Mg supplementation variable (p = 0.01), and therefore confirms the utility of conducting an instrumental variable analysis over logistic regression.

To ensure the use of 2SLS was not invalidated by the non-linearity of the outcome measure, we conducted a second regression using a Probit link function. The results are shown in **Supplementary Table S4**. After accounting for the same measured covariates, Mg supplementation continued to be associated with decreased NOAF (p = 0.01).

## Discussion

In this prescribing preference instrumental variable study, administering supplemental Mg was associated with a 3% decreased chance of developing NOAF, in a general critical care population, after adjusting for observed covariates.

In the context of the two most recent observational studies examining Mg supplementation in the post-cardiac surgery population, the results of this study are contrasting. Both studies found supplemental Mg to be associated with a higher chance of developing post-operative atrial fibrillation ^11, 12^.

Instead, this study concurs with the experimental literature in the same patient population. In a 2019 systematic review and meta-analysis, Chaudhury et al examined 2430 patients, across 20 RCTs of patients undergoing coronary artery bypass surgery. Their results showed Mg supplementation was associated with a significant reduction in post-operative atrial fibrillation (RR 0.76; 95% CI 0.58-0.99; p = 0.04; I2 17.6%). This confirms previous work by Gu et al in the same surgical cohort, but the level of study heterogeneity has been consistently high across all meta-analyses in this field ^10^.

One explanation for the contrasting treatment effect directions seen across observational and experimental work is that the overall treatment effect of Mg supplementation is small, and as such is sensitive to bias. Another explanation of the results from Howitt and Lancaster is confounding by indication. It is possible that using an instrument to correct for unmeasured confounding was able to provide a more reliable estimate of treatment effect, in this case demonstrating that Mg supplementation reduces the risk of NOAF.

In addition to the identification of a more causally robust treatment effect estimate through use of a natural experiment, this study examines this problem in a general adult critical care population. Whilst the importance of NOAF is certainly highest in the post-cardiac surgery population, it is a negative complication in any critically ill patient and thus should be investigated and minimised through the application of evidence-based interventions.

This study has several limitations. It is a single centre study, and so has limited generalisability. Also, there was a large degree of missing data which precluded expanding the study beyond the stated time period. This illustrates some of the difficulties in conducting observational research using routinely collected EHR data. As systems mature and data collection becomes more automated, it is likely this problem will be minimised.

The instrumental variable design itself requires several assumptions which bear mention. First, that levels of the exposure are all adequately represented in the data (stable unit treatment value assumption, SUTVA). Second, that that the instrument (ICU nurse) induces variation in the exposure (administration of Mg). Third, that the instrument may only determine the outcome through its action on the exposure. Fourth, that the instrument operates on an “as-if random” basis, and fifth, that there is no systematic non-compliance with the random nature of the instrument (monotonicity). Except for the second, which is evidenced in the first stage of the analysis, these assumptions are not easily objectively validated. Detailed discussions of the IV assumptions are included in the supplementary materials.

## Conclusions

This study has demonstrated the novel use of a natural experiment, using the CCU nurse’s prescribing preferences as an instrument to define a causally robust estimate of the treatment effect of supplemental Mg for the prophylaxis of NOAF, in a general critical care population. Clinically, this supports the continued administration of supplemental Mg in this context, but further work is necessary to define the optimal target serum Mg for supplementation and define effectiveness in populations with differing baseline risk. This study highlights the problem of continued use of poorly evidenced treatments in critical care medicine. Increasing availability of intelligent EHR systems may help to address these issues, through the efficient and scalable implementation of comparative effectiveness research studies comparing different routine treatment strategies across multiple subgroups.

## Data Availability

Analysis code used in the study is available upon reasonable request to the authors. Enquiries regarding accessing identifiable patient level data may be directed to the UCL Critical Care Health Informatics Collaborative via the corresponding author.

## Acknowledgements

The authors would like to acknowledge the support of the Critical Care Health Informatics Collaborative in generating the data required for this study and Professor Matthew Sydes for reviewing the manuscript. M.G.W. acknowledges funding support from the Medical Research Council, through a Doctoral Training Partnership.

## Declaration of Interests

The authors declare that they have no conflict of interest.

## Funding

MGW is supported by doctoral training program funding via the Medical Research Council.

FWA and SKH are supported by University College London Hospitals National Institute for Health Research Biomedical Research Centre. SKH is supported by a Health Foundation Improvement Science Fellowship.

## Author Contributions

MGW & AR were involved with study design, data preparation, analysis and manuscript drafting.

RK was involved with data preparation and analysis.

SH was involved with study conception, design, analysis and manuscript drafting.

FA was involved with study design and manuscript drafting.

All authors read and approved the final manuscript prior to submission.

## Notes

### Competing Interest Statement

The authors have declared no competing interest.

### Funding Statement

This study did not receive specific funding. The authors received supporting funding as follows:
MGW is supported by doctoral training program funding via the Medical Research Council.
FWA and SKH are supported by University College London Hospitals National Institute for Health Research Biomedical Research Centre. SKH is supported by a Health Foundation Improvement Science Fellowship.

### Author Declarations

This study used routinely collected Electronic Health Record (EHR) data from critical care admissions to a tertiary referral centre in the United Kingdom. Patient data was collected under the auspices of the Critical Care Health Informatics Collaborative (CCHIC). CCHIC has an exemption to collect identifiable clinical data under an opt-out consent framework, for the purposes of conducting secondary research. This is facilitated through an exemption to standard confidentiality law, detailed under Section 251 of the National Health Service (NHS) Act, 2006 (14/CAG/1001) and approved by the National Research Ethics Service (14/LO/103). An application for data access pertinent to the study question was approved by the CCHIC Scientific Advisory Group. Data was stored and analysed in the University College London Data Safe Haven, a secure data environment conforming to NHS Digital's Information Governance Toolkit.

## References

1. Bosch NA, Cimini J, Walkey AJ. Atrial Fibrillation in the ICU. Chest 2018; 154: 1424–34

2. Burrage PS, Low YH, Campbell NG, O’Brien B. New-Onset Atrial Fibrillation in Adult Patients After Cardiac Surgery. Curr Anesthesiol Rep 2019; 9: 174–93

3. Amar D. Postthoracotomy atrial fibrillation: Current Opinion in Anaesthesiology 2007; 20: 43–7

4. Stawicki SPA, Prosciak MP, Gerlach AT, et al. Atrial fibrillation after esophagectomy: an indicator of postoperative morbidity. Gen Thorac Cardiovasc Surg 2011; 59: 399–405

5. Chebbout R, Heywood EG, Drake TM, et al. A systematic review of the incidence of and risk factors for postoperative atrial fibrillation following general surgery. Anaesthesia 2018; 73: 490–8

6. Moss TJ, Calland JF, Enfield KB, et al. New-Onset Atrial Fibrillation in the Critically Ill*. Critical Care Medicine 2017; 45: 790–7

7. Rajagopalan B, Shah Z, Narasimha D, et al. Efficacy of Intravenous Magnesium in Facilitating Cardioversion of Atrial Fibrillation. Circ Arrhythm Electrophysiol [Internet] 2016 [cited 2020 Jun 7]; 9 Available from: https://www.ahajournals.org/doi/10.1161/CIRCEP.116.003968

8. Garg J, Chaudhary R, Krishnamoorthy P, Shah N, Sharma A, Bozorgnia B. ROLE OF PROPHYLACTIC MAGNESIUM SUPPLEMENTATION IN PREVENTION OF POSTOPERATIVE ATRIAL FIBRILLATION IN PATIENTS UNDERGOING CORONARY ARTERY BYPASS GRAFTING: A META-ANALYSIS OF 23 RANDOMIZED CONTROLLED TRIALS. Journal of the American College of Cardiology 2016; 67: 689

9. Arsenault KA, Yusuf AM, Crystal E, et al. Interventions for preventing post-operative atrial fibrillation in patients undergoing heart surgery. Cochrane Heart Group, editor. Cochrane Database of Systematic Reviews [Internet] 2013 [cited 2020 Jun 9]; Available from: http://doi.wiley.com/10.1002/14651858.CD003611.pub3

10. Gu W-J, Wu Z-J, Wang P-F, Aung LHH, Yin R-X. Intravenous magnesium prevents atrial fibrillation after coronary artery bypass grafting: a meta-analysis of 7 double-blind, placebo-controlled, randomized clinical trials. Trials 2012; 13: 41

11. Howitt SH, Grant SW, Campbell NG, Malagon I, McCollum C. Are Serum Potassium and Magnesium Levels Associated with Atrial Fibrillation After Cardiac Surgery? Journal of Cardiothoracic and Vascular Anesthesia 2020; 34: 1152–9

12. Lancaster TS, Schill MR, Greenberg JW, et al. Potassium and Magnesium Supplementation Do Not Protect Against Atrial Fibrillation After Cardiac Operation: A Time-Matched Analysis. The Annals of Thoracic Surgery 2016; 102: 1181–8

13. Naksuk N, Hu T, Krittanawong C, et al. Association of Serum Magnesium on Mortality in Patients Admitted to the Intensive Cardiac Care Unit. The American Journal of Medicine 2017; 130: 229.e5-229.e13

14. Brookhart MA, Schneeweiss S. Preference-based instrumental variable methods for the estimation of treatment effects: assessing validity and interpreting results. Int J Biostat 2007; 3: 14

15. Keele L, Sharoky CE, Sellers MM, Wirtalla CJ, Kelz RR. An Instrumental Variables Design for the Effect of Emergency General Surgery. Epidemiologic Methods [Internet] 2018 [cited 2020 Jun 6]; 7 Available from: http://www.degruyter.com/view/j/em.2018.7.issue-1/em-2017-0012/em-2017-0012.xml

16. Harris S, Shi S, Brealey D, et al. Critical Care Health Informatics Collaborative (CCHIC): Data, tools and methods for reproducible research: A multi-centre UK intensive care database. International Journal of Medical Informatics 2018; 112: 82–9

17. Staiger DO, Stock JH. Instrumental Variables Regression with Weak Instruments. Econometrica 1997; 65: 557–86

18. Hausman JA. Specification Tests in Econometrics. Econometrica 1978; 46: 1251

19. Core Team R. R: A language and environment for statistical computing. [Internet]. Vienna, Austria: R Foundation for Statistical Computing; 2020. Available from: https://www.R-project.org/

20. StataCorp. Stata Statistical Software: Release 16. StataCorp LLC; 2019.

