## Supplementary Materials for "Prophylactic Magnesium Supplementation and New-Onset Atrial Fibrillation in a General Critical Care Population: A Prescribing Preference Instrumental Variable Analysis"

### Methods

#### S1 - Data and Transformations

| **Variable** | **Units** | **Transformation** |
| --- | --- | --- |
| Age | Years, Continuous |  |
| Sex | Female \| Male, Categorical |  |
| Length of ICU Stay | Days, Continuous |  |
| Acute Physiology And Chronic Health Evaluation (APACHE II) Score (on admission) | Numerical, Continuous | Standardised |
| ICU Mortality | Yes \| No, Categorical |  |
| Serum Magnesium | Millimoles per Litre, Continuous | Standardised |
| Serum Potassium | Millimoles per Litre, Continuous | Standardised |
| Serum pH | -log[H^+^], Continuous | Standardised |
| Magnesium Administration | Yes \| No, Categorical |  |
| Heart Rhythm | Sinus Rhythm & Variants \| Atrial Fibrillation & Variants, Categorical |  |
| Heart Rate | Beats per Minute, Continuous | Standardised |
| Noradrenaline | Micrograms per Kilogram per Minute, Continuous | Standardised |

#### S2 - Example of Magnesium Observation Windows

| **Patient Identity Number** | **Episode Identity Number** | **Serum Mg Laboratory Timestamp** | **Serum Mg Level** | **Next Serum Mg Laboratory Timestamp** | **24 Hour post initial Serum Mg Timestamp** | **Episode End Timestamp** | **Mg Window Censor Timestamp** |
| --- | --- | --- | --- | --- | --- | --- | --- |
| 1 | A | 1.1.19 0800 | 1.20 | 2.1.19 0800 | 2.1.19 0800 | 14.1.19 2358 | 2.1.19 0600 |
| 1 | A | 2.1.19 0600 | 0.80 | NA | 3.1.19 0600 | 14.1.19 2358 | 3.1.19 0600 |
| 2 | A | 14.12.19 0753 | 0.79 | 14.12.19 0000 | 15.12.19 0853 | 15.12.19 0853 | 14.12.19 2000 |

### Results

#### S3 - Effect of Nurse on Magnesium Administration

| **Variable** | **Estimate** | **Standard Error** | **P value** |
| --- | --- | --- | --- |
| Intercept | -0.93 | 0.05 | < 0.001 |
| Serum Magnesium (std.) | -2.08 | 0.05 | < 0.001 |
| Heart Rate (std.) | 0.19 | 0.03 | < 0.001 |
| pH (std.) | 0.17 | 0.03 | < 0.001 |
| Prev. AF | 0.36 | 0.08 | < 0.001 |
| APACHE (std.) | 0.10 | 0.03 | < 0.001 |
| Mean Noradrenaline Prev. 24 hours | 0.13 | 0.03 | < 0.001 |

#### S4 - Effect of Magnesium Supplementation on New-Onset Atrial Fibrillation, Probit Model

| **Variable** | **Estimate** | **Standard Error** | **P value** |
| --- | --- | --- | --- |
| Intercept | -1.7988 | 0.265 | < 0.001 |
| Mg Supplementation | -0.7966 | 0.314 | 0.011 |
| Age (std.) | 0.3097 | 0.048 | < 0.001 |
| Sex (Male) | -0.1297 | 0.068 | 0.058 |
| APACHE (std.) | -0.0019 | 0.368 | 0.959 |
| Prev. AF | 0.6782 | 0.726 | < 0.001 |
| Serum Magnesium (std.) | -0.0868 | 0.094 | 0.354 |
| Serum Potassium (std.) | 0.0123 | 0.036 | 0.730 |
| Heart Rate (std.) | 0.1221 | 0.036 | 0.001 |
| pH (std.) | 0.0378 | 0.035 | 0.280 |
| Mean Noradrenaline Prev. 24 hours | 0.1170 | 0.266 | < 0.001 |

#### Marginal Effects Modelling for Probit Model

We calculated the Average Treatment Effect (ATE) of magnesium supplementation on incidence of AF based on results from the IV probit regression. We used 2 methods of calculation 1. The `margins, dydx(*) predict(pr)` postestimation command in Stata, which estimated an ATE of 0.043. 2. We calculated the difference between predicted incidence of AF when all windows were assumed to have been supplemented with magnesium, compared with all windows not being supplemented with magnesium. This method yielded an estimated ATE of 0.034. Both these estimates are approximately consistent with the estimated effect from the 2 stage least squares linear regression which was 0.033.

### Discussion

#### Instrumental Variable Assumptions

In exploring IV assumptions we refer to work done by Keele et al, exploring surgeon preference as an instrumental variable for the effect of emergency surgery^24^.

##### Stable Unit Treatment Value Assumption

This assumption asserts that all possible levels of the exposure are represented in the data. In this study we examined Mg administration as a binary exposure. It is possible therefore that this assumption is violated if the doses of Mg administered varied e.g. 2g vs. 5g. Given the ‘as required’ prescription was electronic and pre-formatted, the variance of dose administered may be assumed to be small. In addition, we have not distinguished between oral and intravenous Mg supplementation. Despite the good bioavailability of oral Mg, it is reasonable to presume that any protective effect of oral supplementation might be slower than intravenous and potentially smaller overall. It is also possible that some patients received multiple independent administrations of Mg within one treatment window.

The second component to this assumption is that it is not possible to affect an individual patient’s outcome through other patient’s exposures. This may be a relevant concern in resource limited settings where allowing one patient treatment might preclude the same treatment being available for another patient. In this setting, each patient is allocated an individual ICU nurse and there is no resource limitation for supplemental Mg.

##### Instrument Affects Exposure

This assumption requires that the instrument (nurse) induces variation in the exposure (Mg administration). In this study we have demonstrated this through the creation of a multi-level model to predict Mg administration. This suggests that individual nurse preference has a significant role in determining eventual Mg administration after accounting for the covariates included. This can be confirmed by observing the predicted probabilities of Mg administration.

##### Exclusion Restriction

The exclusion restriction mandates that the instrument (nurse) cannot affect the outcome (AF), other than directly through its action on the exposure (Mg administration). In this study, having the outcome is directly and indirectly influenced by a multitude of factors, some of which potentially lie within the control of the ICU nurse, and as such may violate this assumption. One such example is the supplementation of Potassium. Whilst we accounted for baseline serum Potassium concentrations, we did not account for Potassium supplementation by the ICU nurse. If supplementation of Potassium was either systematically conducted or withheld, then this may induce bias in the study result.

##### As-If Random Instrument

This assumption mandates that the application of the instrument to the patient is genuinely random in nature. This is difficult to reliably extrapolate from the data, but would conceivably require the systematic application of more experienced ICU nurses (with pre-set beliefs about the utility of Mg supplementation) to sicker patients. It is possible to partially evaluate this through examining the balance of ‘Pro’ and ‘Anti’ Mg groups with respect to patient characteristics. As illustrated in Table 1 of the main text, the groups appear well balanced in all observed characteristics with the exception of frequency of Mg administration. This of course does not exclude group imbalance in potentially important covariates which are unobserved.

##### Monotonicity

The final assumption requires that there is no systematic non-compliance with the “as-if random” application of the instrument. In this case, this would require nurses to systematically administer Mg when serum levels are high and vice versa. This is counterintuitive and unlikely.

It is possible that this assumption may be violated if there is a group of patients for which nurses systematically aim for higher serum Mg levels in (e.g. pre-existing AF). In these cases, a high level of the IV (high serum Mg) may not preclude the administration of additional Mg, in the quest for an even higher target in this subgroup.
